# Preliminary findings from the Dynamics of the Immune Responses to Repeat Influenza Vaccination Exposures (DRIVE I) Study: a Randomized Controlled Trial

**DOI:** 10.1101/2024.05.16.24307455

**Authors:** Benjamin J. Cowling, Sook-San Wong, Jefferson J. S. Santos, Lisa Touyon, Jordan Ort, Naiqing Ye, Natalie K. M. Kwok, Faith Ho, Samuel M. S. Cheng, Dennis K. M. Ip, Malik Peiris, Richard J. Webby, Patrick C. Wilson, Sophie A. Valkenburg, John S. Tsang, Nancy H. L. Leung, Scott E. Hensley, Sarah Cobey

## Abstract

**Background:** Studies have reported that repeated annual vaccination may influence the effectiveness of the influenza vaccination in the current season. The mechanisms underlying these differences are unclear but might include “focusing” of the adaptive immune response to older strains.

**Methods:** We established a 5-year randomized placebo-controlled trial of repeated influenza vaccination (Flublok, Sanofi Pasteur) in adults 18-45 years of age. Participants were randomized equally between five groups, with planned annual receipt of vaccination (V) or saline placebo (P) as follows: P-P-P-P-V, P-P-P-V-V, P-P-V-V-V, P-V-V-V-V, or V-V-V-V-V. Serum samples were collected each year just before vaccination and after 30 and 182 days. A subset of sera were tested by hemagglutination inhibition assays, focus reduction neutralization tests and enzyme-linked immunosorbent assays against vaccine strains.

**Results:** From 23 October 2020 through 11 March 2021 we enrolled and randomized 447 adults. We selected sera from 95 participants at five timepoints from the first two study years for testing. Among vaccinated individuals, antibody titers increased between days 0 and 30 against each of the vaccine strains, with substantial increases for first-time vaccinees and smaller increases for repeat vaccinees, who had higher pre-vaccination titers in year 2. There were statistically significant reductions in the proportion of participants achieving a four-fold greater rise in antibody titer for the repeat vaccinees for A(H1N1), B/Victoria and B/Yamagata, but not for influenza A(H3N2). There were no statistically significant differences between groups in geometric mean titers at day 30 or the proportions of participants with antibody titers ≥40 at day 30 for any of the vaccine strains.

**Conclusions:** In the first two years, repeat vaccinees and first-time vaccinees had similar post-vaccination geometric mean titers to all four vaccine strains, indicative of similar levels of clinical protection. The vaccine strains of A(H1N1) and A(H3N2) were updated in year 2, providing an opportunity to explore antigenic distances between those strains in humans in subsequent years.

## INTRODUCTION

The efficacy of influenza vaccines has been demonstrated in randomized trials, and annual influenza vaccination campaigns prevent considerable morbidity and mortality [1]. Continuous viral evolution necessitates regular updates to vaccine strains, and the degree of match between vaccine and circulating strains affects vaccine protection [2]. A number of studies have reported that repeated annual vaccination may influence the effectiveness of the influenza vaccination in the current season [3–7]. The effect of repeated vaccination on immunogenicity has been less frequently assessed in multi-year randomized trials, but repeat vaccination effects are also observed [8–12]. The mechanisms underlying these differences are unclear, but might include “focusing” of the adaptive immune response to older strains [13]. Under this model, repeated exposures boost responses to conserved and potentially less protective epitopes, which might in some years reduce protection against circulating strains [14, 15].

Enhanced influenza vaccines, including the recombinant hemagglutinin vaccine Flublok (Sanofi Pasteur), stimulate stronger immune responses and may be able to overcome repeat vaccination effects. The Flublok vaccine has two major differences with the standard egg-grown inactivated influenza vaccine [10, 16, 17]. First, because eggs are not used in the production process for Flublok, the antigens included in the vaccine are more similar to circulating viruses, circumventing the issue of egg-adapted mutations in the hemagglutinin (HA) protein that can lead to antigenic mismatch [10, 18]. Second, it includes three times more HA antigen than standard-dose vaccines and can therefore generate a stronger, more HA-specific humoral immune response [10, 11]. Flublok has been approved by the Food and Drug Administration for use in the United States in all adults ≥18 years of age since October 2014 [19]. Evidence from randomized trials have shown improved immunogenicity and efficacy and a slightly lower local reactogenicity compared with standard inactivated influenza vaccine in adults ≥18 years [20].

We designed a randomized controlled trial to explore immune responses to first-time or repeated influenza vaccination with the Flublok vaccine, with a particular interest in the possible occurrence of repeat vaccination effects.

## METHODS

### Study design

This study is a randomized controlled trial in adults 18-45 years of age at enrolment (Clinicaltrials.gov: NCT04576377). Participants were enrolled from the general community in Hong Kong, with study advertisements distributed via institutions (such as schools and universities), organizations (such as professional associations), and local community centers, and through mass promotion efforts including mass mailing to residential estates, advertisements in newspapers and public transport, social media platforms (such as Facebook), bulk emails, and invitation to and referrals from members of previous studies. Individuals were eligible to participate if they were between 18 and 45 years of age, capable of providing informed consent, and intending to reside in Hong Kong for at least the next two years. Potential participants were excluded if they had been vaccinated against influenza in the preceding 24 months, if they were included in a priority group for influenza vaccination (e.g. healthcare worker, pregnant woman), if they had a diagnosed immunosuppressive condition or were taking immunosuppressive medication, or if they had severe allergies or bleeding conditions that contraindicated intramuscular influenza vaccination.

We collected a baseline 9-ml clotted blood sample from enrolled participants, and we used a standardized questionnaire to collect baseline information on demographics, current health status, medical history and medication use. Participants were randomized equally among five equal groups using a permuted block approach with block sizes of 5 and 10, using the statistical software package R. Based on the randomization scheme (Appendix Figure 1), each participant would receive either influenza vaccination or placebo annually in the following schedule for the first four years of the trial in the following schedule: Group 1, placebo injection for all four years; Group 2, placebo for three years followed by influenza vaccination in the fourth year; Group 3, placebo for two years followed by vaccination for two years; Group 4, placebo in the first year and then vaccination for the subsequent three years; and Group 5, vaccination in all four years. All participants would then receive vaccination in the fifth and final year. Allocation to these groups was concealed using REDCap [21].

The influenza vaccine used in our trial is recombinant HA quadrivalent influenza vaccine (0.5mL Flublok®, Sanofi Pasteur) for the northern hemisphere. For the 2020/21 northern hemisphere influenza season, it was formulated to contain 180 mg HA per 0.5-mL dose, with 45 mg HA of each of the following four influenza virus strains: A/Hawaii/70/2019 (H1N1), A/Minnesota/41/2019 (an A/Hong Kong/45/2019-like virus) (H3N2), B/Washington/02/2019 and B/Phuket/3073/2013. In 2021/22 the influenza B strains were unchanged while the influenza A strains were updated to A/Wisconsin/588/2019 (H1N1) and A/Tasmania/503/2020 (an A/Cambodia/e0826360/2020-like virus) (H3N2) (Appendix Table 1). To maintain blinding of participants, we used 0.5 ml saline placebo in syringes prepared in advance and packed the vaccines and placebo doses in numbered boxes according to the randomization scheme. Other than the nurse who conducted the injections, all other study staff were blinded to the study intervention.

Following vaccination, we invited participants to return for scheduled follow-up visits at 30 days and 182 days after vaccination for further blood draws. In a subset of participants we arranged for collection of peripheral blood mononuclear cells at baseline, day 7 and day 30, and additional clotted blood samples on days 91 and 273. All participants were invited to report any acute respiratory illnesses. During periods of influenza activity, we planned to implement weekly active illness surveillance, collecting nasal swabs from ill participants for testing by PCR and an additional blood sample 30 days after illness onset.

### Ethics

Written informed consent was obtained from all participants. Participants were compensated with a gift voucher worth HK$100 (US$13) at each blood draw. The study protocol was approved by the Institutional Review Boards of the University of Hong Kong (ref: UW19-551) and of the University of Chicago Biological Sciences Division (ref: IRB20-0217).

### Primary and secondary outcomes

The primary outcome measure in this trial is the vaccine immunogenicity, measured in terms of antibody titers in hemagglutination-inhibition (HAI) assays or foci reduction neutralization tests (FRNTs) for each vaccine strain. We compared the proportion of participants who achieved the targeted rise in antibody titre against each of the vaccine strains at 30 days. The targeted rise in antibody titre is defined as a four-fold or greater rise in titer, including either a pre-vaccination HAI titer <10 and a post-vaccination HAI titre ≥20 or a pre-vaccination HAI titer ≥10 and at least a four-fold rise in post-vaccination HAI antibody titer. We also assessed the geometric mean titer (GMT) ratios between the various randomized groups against each of the vaccine strains at 30 days and 182 days. HAI assays were completed with A(H1N1) and influenza B viruses; however; we used FRNTs to quantify antibodies against A(H3N2) strains, since some contemporary A(H3N2) strains inefficiently agglutinate red blood cells [10, 22]. Since the FRNT dilution series started at 20 instead of 10 due to the assay design, we defined a targeted rise for antibody titers against A(H3N2) measured by FRNT as a 4-fold rise to a post-vaccination titer of ≥40.

Other secondary outcomes specified in the protocol include additional comparisons of antibody titers; analyses of cellular immunity, including transcriptional activity of immune cells; comparisons of adverse reactions after vaccination; and the occurrence of influenza or other acute respiratory illnesses. These secondary outcomes will be addressed in subsequent reports. Of particular note, there was no influenza circulation in Hong Kong between March 2020 and February 2023, which included the first two years of the trial [23]. Only a few influenza virus infections were detected by the local public health laboratory during this period, and they were mostly in arriving travelers and children shedding live attenuated vaccine virus [24, 25].

### Laboratory analysis

Blood samples were collected in tubes for clotted blood, stored in a refrigerated container at 2-8°C immediately and delivered to the laboratory within two days for further processing. Serum specimens were aliquoted and stored at -80°C prior to subsequent testing. Serum samples were treated with receptor destroying enzyme (RDE) and tested by HAI against the influenza A(H1N1) and B vaccine strains using a standard protocol [26]. For influenza A(H1N1), the vaccine strains were A/Hawaii/70/2019 in 2020/21 (GISAID Accession # EPI397028) and A/Wisconsin/588/2019 (GISAID Accession # EPI404460) in 2021/22. Due to the inability to obtain sufficient volume of cell-grown virus stock, A/Wisconsin/588/2019 was eventually propagated once in eggs. A single mutation D204V was found in the egg-grown stock. A comparison of antigenicity using 22 serum samples from five participants showed that assay with the egg-grown stock had slightly higher sensitivity that yielded 2-fold higher titers in 50% of the samples but otherwise showed good correlation with cell-grown stock (Appendix Figure 2). For influenza B, ether-treated egg-grown antigens were used, and the vaccine strains were B/Washington/02/2019 (GISAID Accession # EPI347829) and B/Phuket/3073/2014 (GISAID Accession # EPI168822) in both years. We used a focus reduction neutralization test (FRNT) for the two influenza A(H3N2) vaccine strains A/Hong Kong/45/2019 in 2020/21 and A/Cambodia/e0826360/2020 in 2021/22. A/Hong Kong/45/2019 HA (GISAID Accession # EPI1397376) is identical at the amino acid level to A/Minnesota/41/2019 HA (GISAID Accession # EPI1487157) included in the Flublok for 2020/21 influenza season. However, A/Cambodia/e0826360/2020 HA (GISAID Accession # EPI1837753) differs by a single amino acid substitution (N171K at antigenic site D) from A/Tasmania/503/2020 HA (GISAID Accession # EPI1759269) included in the Flublok for 2021/22 influenza season. Both influenza A(H3N2) virus strains were generated by reverse genetics using A/Puerto Rico/8/1934 internal genes [27]. FRNT assays were completed as previously described [28].

To estimate total binding antibody of specific IgG responses, Enzyme Linked Immunosorbent Assays (ELISAs) were performed with recombinant HA (rHA) of the A(H1N1) vaccine strains A/Hawaii/70/2019 in 2020/21 and A/Wisconsin/588/2019, and the A(H3N2) vaccine strains A/Minnesota/41/2019 and A/Tasmania/503/2020 (Appendix Table 1). The HA stalk monoclonal antibody CR9114 was used as an internal control on each plate. Expression and purification of rHAs and CR9114 was performed as previously described [29].

### Sample size justification

We aimed to enroll 820 participants into our study. This would permit us to have a sample size of at least 100 participants in each of the five groups at the fourth year of follow-up, allowing for an anticipated drop-out rate of 15% per year without replacement. Most of our outcome measures, including both of our primary outcome measures, are based on geometric mean antibody titers. In year 2, with a target sample size of 139 in each group, we expected to have 80% power to identify 1.6-fold differences in GMT between groups, assuming a standard deviation of log_2_(GMT) of 1.8. However, due to disruption in study activities during the COVID-19 pandemic we were unable to reach our target sample size, and as a consequence we updated the protocol to include enrolment of an additional cohort of 530 participants starting in 2021/22 with a similar trial design, with participants randomized across four groups instead of five, and receiving four annual doses of vaccination/placebo as part of the study instead of five. Results from the second cohort with participant enrolment in 2021/22, named DRIVE II, will be reported in due course. For consistency, the present cohort with participant enrolment in 2020/21 is named DRIVE I.

### Statistical analysis

We assessed the proportion of participants who achieved a 4-fold or greater rise in antibody titre against each of the vaccine strains at 30 days, and compared these proportions between the vaccine-vaccine, placebo-vaccine and placebo-placebo groups using Fisher exact tests, pooling the three placebo-placebo groups together. We estimated GMT ratios versus the placebo group at day 30 of year 2 and compared them between first-time vaccines and repeat vaccinees using t-tests on the log-transformed GMT ratios. Confidence intervals were estimated using t distributions. All analyses were conducted in R version 4.2.1 (R Foundation for Statistical Computing, Vienna, Austria). Data and R syntax will be made available after publication.

## RESULTS

From 23 October 2020 through 11 March 2021, 447 individuals were enrolled and received influenza vaccination or placebo according to the randomization scheme, with 88 to 91 individuals in each arm (Figure 1). The median age of participants was 31 years, and 54% were male, with similar characteristics across the five arms (Table 1). None of the participants reported receiving influenza vaccination in the two years prior to the start of the trial, and only 12% reported ever previously receiving influenza vaccination. We randomly selected for laboratory analysis a subset of 15 participants from groups 1-3 (who received placebo in both years), 40 participants from group 4 (placebo followed by vaccination), and 40 participants from group 5 (vaccination in both years), and noted similar characteristics in this subset to the overall participants (Appendix Table 2).

**Figure 1.**
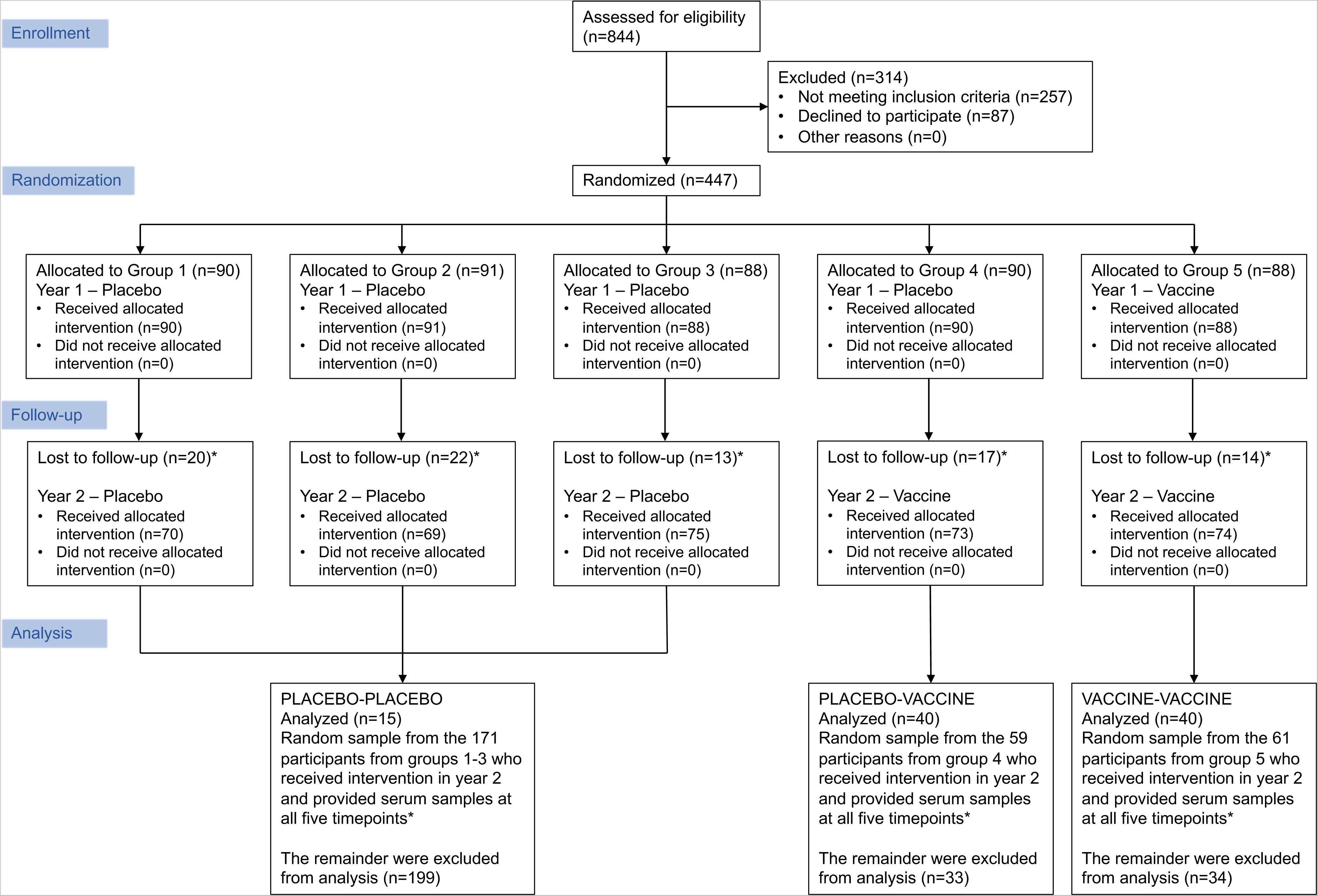
Study flow chart showing participant enrolment into the study, randomization into five groups, interventions received, and follow up. Footnote: *The random samples were selected from participants who provided serum samples at these five timepoints: year 1 day 0, day 30 and day 182, and year 2 day 0 and day 30.

**Table 1.** Characteristics of participants randomized to different vaccination strategies.

| Characteristic | Randomized allocation to receipt of placebo (P) or influenza vaccination (V)<br>in five consecutive years |  |  |  |  |
| --- | --- | --- | --- | --- | --- |
|  | Group 1<br>P-P-P-P-V<br>(n=90) | Group 2<br>P-P-P-V-V<br>(n=91) | Group 3<br>P-P-V-V-V<br>(n=88) | Group 4<br>P-V-V-V-V<br>(n=90) | Group 5<br>V-V-V-V-V<br>(n=88) |
| Age at randomization |  |  |  |  |  |
| 18-25 years | 27 (30%) | 27 (30%) | 34 (39%) | 22 (24%) | 20 (23%) |
| 26-35 years | 32 (36%) | 32 (35%) | 28 (32%) | 34 (38%) | 34 (39%) |
| 36-45 years | 31 (34%) | 32 (35%) | 26 (30%) | 34 (38%) | 34 (39%) |
| Male sex | 48 (53%) | 60 (66%) | 42 (48%) | 46 (51%) | 44 (50%) |
| Reported receipt of influenza vaccination in the two years prior to the trial | 0 (0%) | 0 (0%) | 0 (0%) | 0 (0%) | 0 (0%) |
| Reported ever receiving influenza vaccination prior to the trial | 11 (12%) | 11 (12%) | 15 (17%) | 9 (10%) | 9 (10%) |

Antibody titers increased between days 0 and 30 against each of the vaccine strains, with greater fold increases for first-time vaccinees compared to repeat vaccinees, while the repeat vaccinees had higher antibody titers prior to vaccination in year 2 (Figure 2). At day 30 of year 2, the GMTs were similar in the first-time vaccinees and repeat vaccinees (groups 4 and 5, respectively) for each strain analyzed (Appendix Table 4). While there were statistically significant reductions in the proportion achieving a four-fold greater rise in antibody titer for the repeat vaccinees in year 2 for A(H1N1), B/Victoria and B/Yamagata, there were no statistically significant differences in GMTs at day 30 or the proportions with antibody titers ≥40 at day 30 for any of the strains (Table 2).

**Figure 2.**
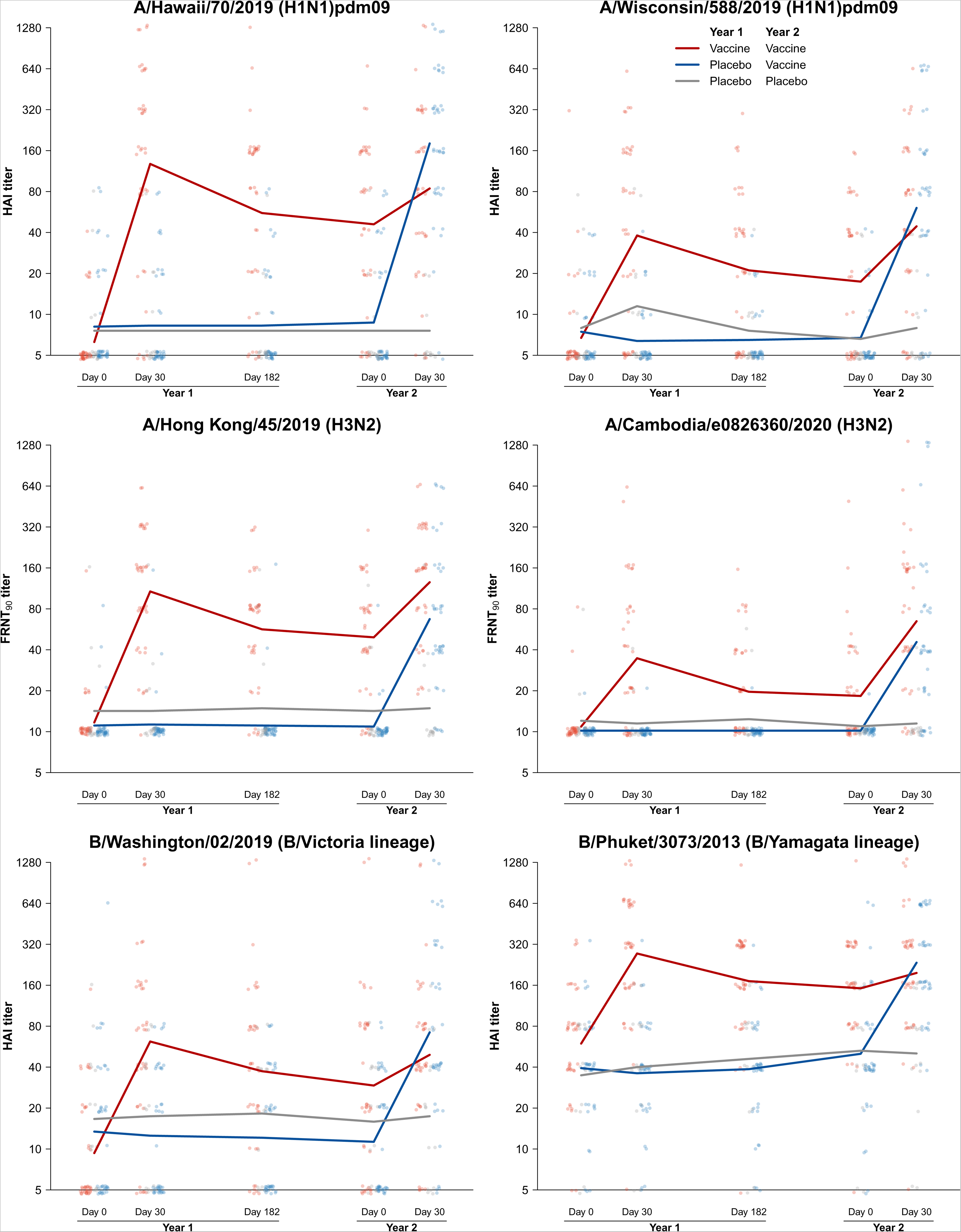
Antibody titers at various timepoints measured by hemagglutination inhibition assay for influenza A(H1N1) and B, and by focus reduction neutralization test for influenza A(H3N2). Measured titers are plotted for each group at 0, 30 and 182 days post-vaccination of year 1 and at 0 and 30 days post-vaccination of year 2, and lines represent the geometric mean titers at each timepoint. Data from group 5, receiving vaccine in both years, are shown in red. Data from group 4, receiving placebo in year 1 and vaccine in year 2, are shown in blue. Data from the other groups, receiving placebo in both years, are shown in gray.

**Table 2.**
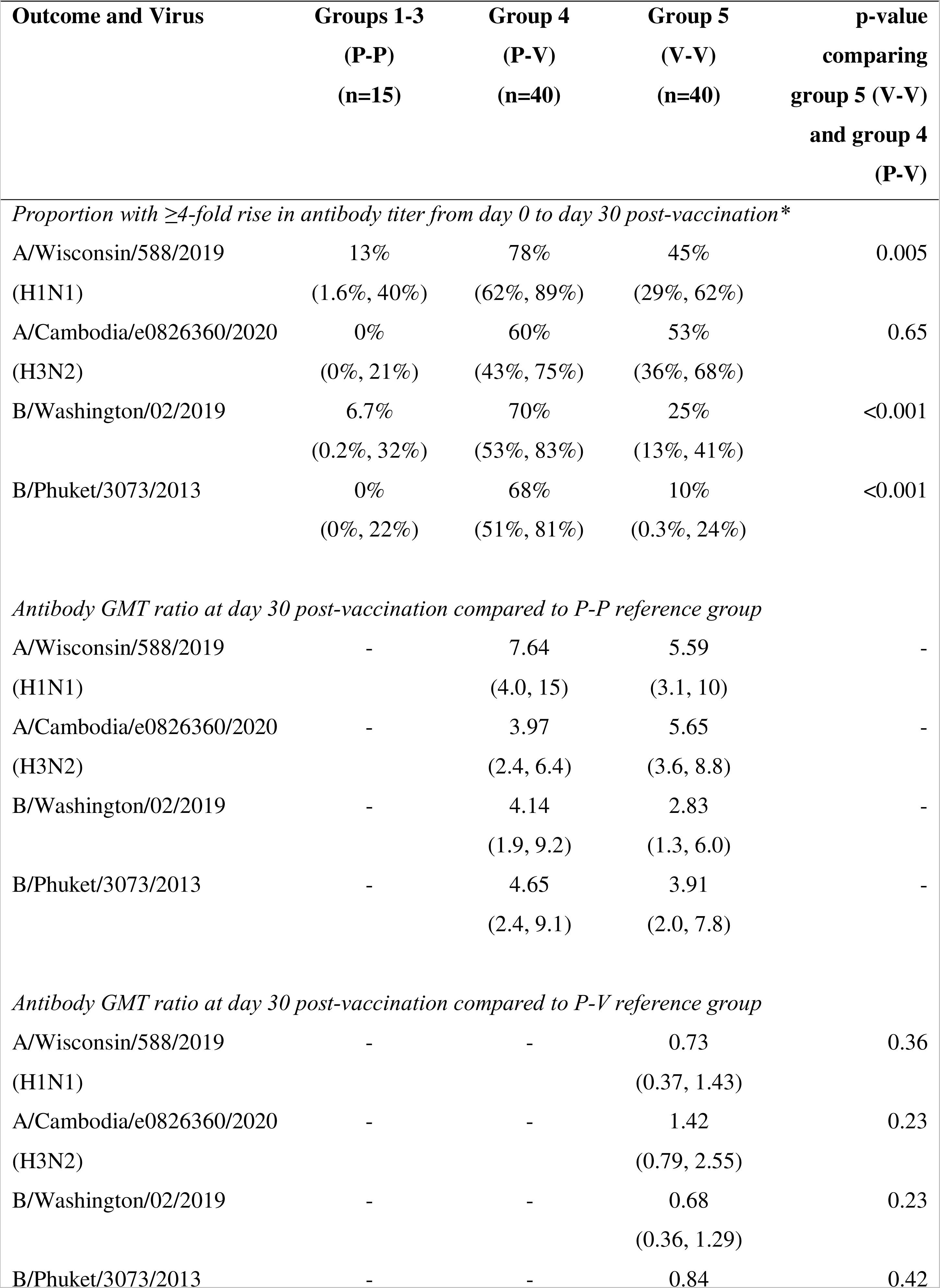
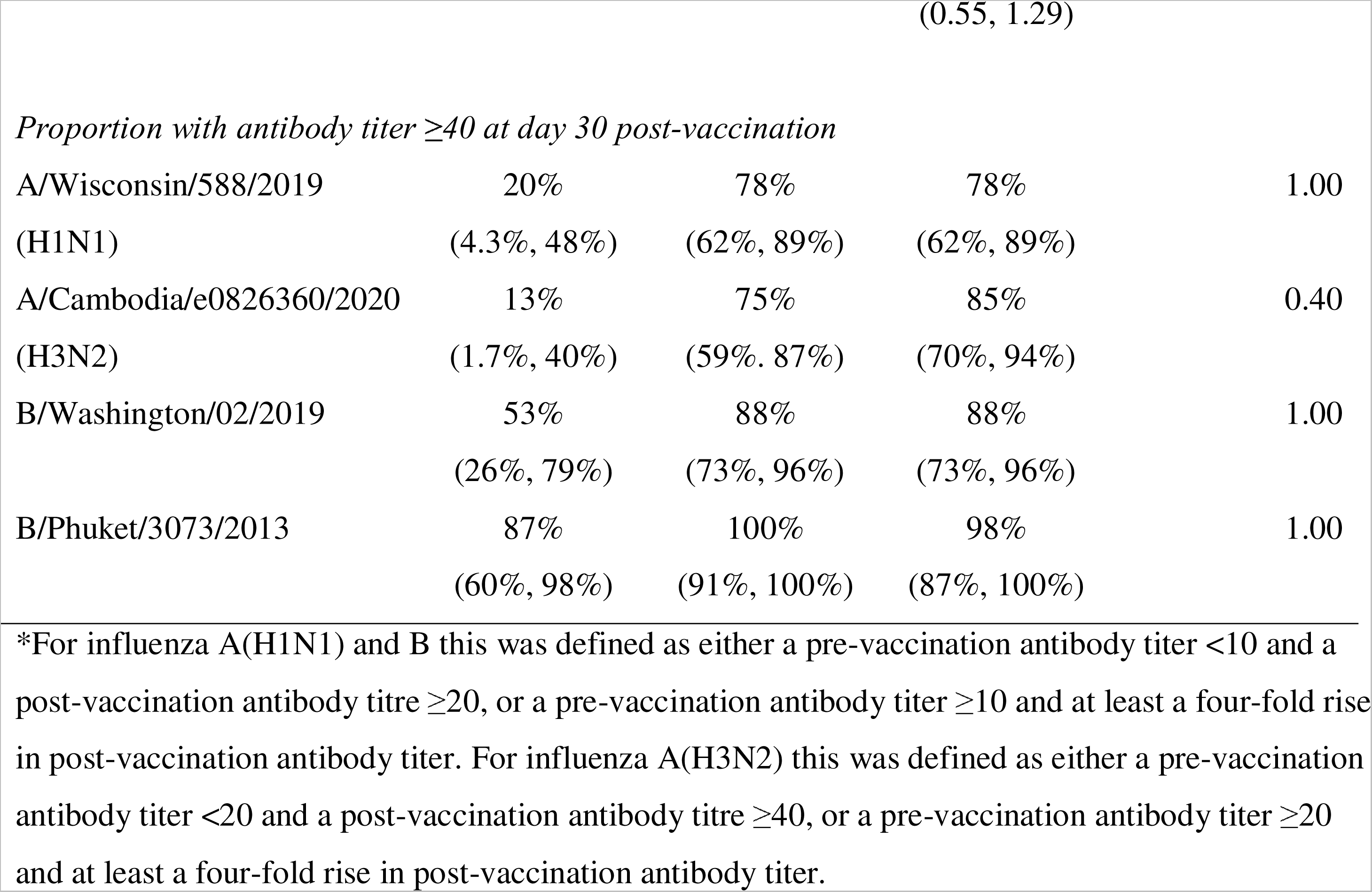
Antibody responses to influenza vaccination and placebo in year 2 of the trial, with 95% confidence intervals. Antibody responses were measured by HAI for influenza A(H1N1) and B, and by FRNT for influenza A(H3N2).

The A(H1N1) and A(H3N2) components of the northern hemisphere vaccine were updated between 2020/21 and 2021/22. For each subtype, we examined whether vaccination with one strain increased antibody titers to the other, which is a measure of cross-reactivity or antigenic distance. We focused on the responses in first-time vaccinees in each of the two years (i.e. group 5 in year 1 and group 4 in year 2). For A(H1N1), the year 1 GMT at day 30 was approximately 3.2-fold higher to the 2020/21 vaccine strain (A/Hawaii/70/2019) than to the vaccine strain used the next year (A/Wisconsin/588/2019), consistent with a modest antigenic distance between the two strains (Table 3). For A(H3N2), the fold-difference between the day 30 GMT to the 2020/21 strain (A/Hong Kong/45/2019) and the 2021/22 strain (A/Cambodia/e0826360/2020) was approximately 3.1, also indicating modest antigenic distance.

**Table 3.**
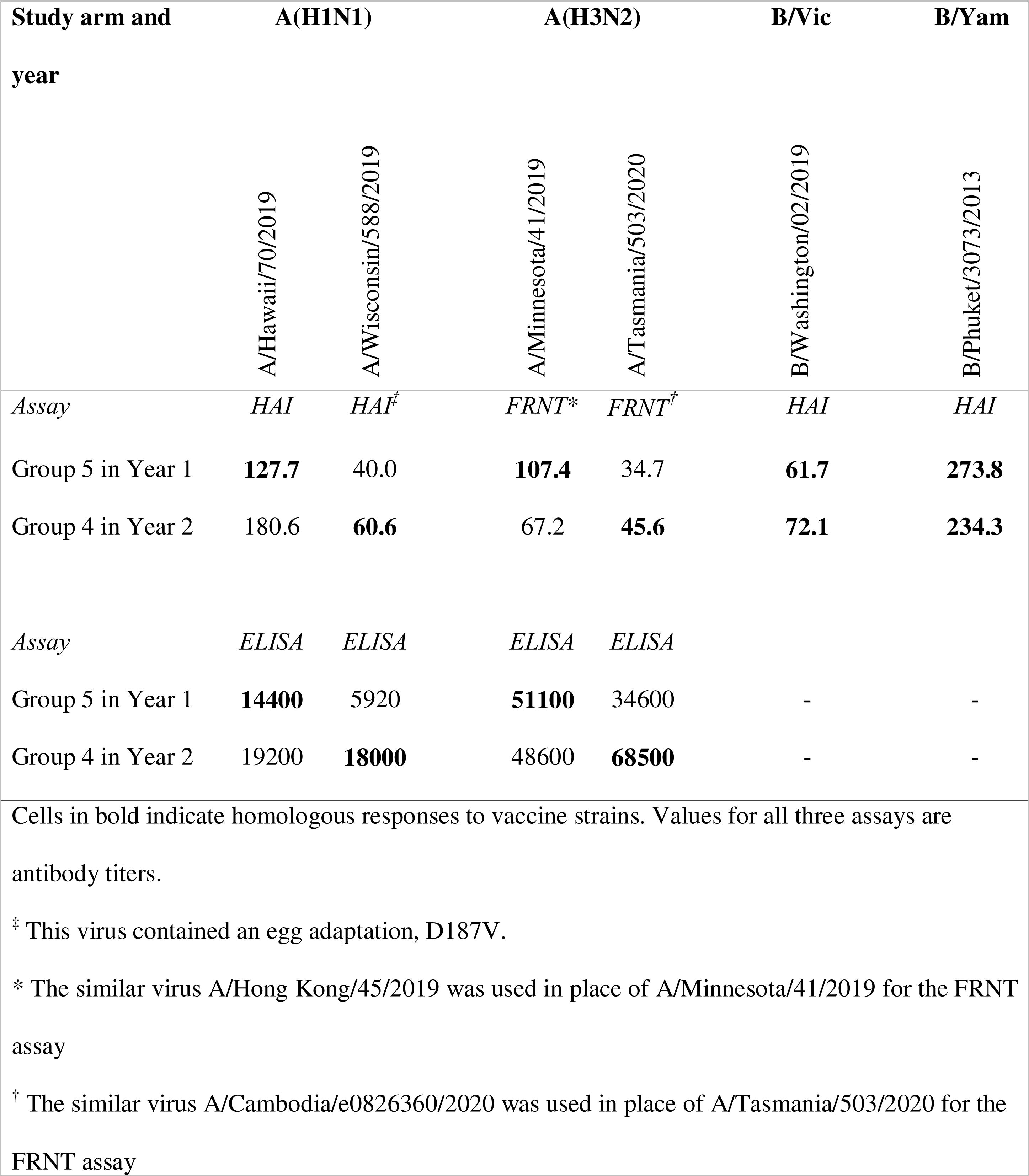
Geometric mean antibody levels 30 days post-vaccination for participants receiving vaccination for the first time in the study.

| Study arm and year | A(H1N1) |  | A(H3N2) |  | B/Vic | B/Yam |
| --- | --- | --- | --- | --- | --- | --- |
|  | A/Hawaii/70/2019 | A/Wisconsin/588/2019 | A/Minnesota/41/2019 | A/Tasmania/503/2020 | B/Washington/02/2019 | B/Phuket/3073/2013 |
| Assay | *HAI* | *HAI*<sup>‡</sup> | *FRNT*<sup>*</sup> | *FRNT*<sup>†</sup> | *HAI* | *HAI* |
| Group 5 in Year 1 | **127.7** | 40.0 | **107.4** | 34.7 | **61.7** | **273.8** |
| Group 4 in Year 2 | 180.6 | **60.6** | 67.2 | **45.6** | **72.1** | **234.3** |
| Assay | *ELISA* | *ELISA* | *ELISA* | *ELISA* |  |  |
| Group 5 in Year 1 | **14400** | 5920 | **51100** | 34600 | - | - |
| Group 4 in Year 2 | 19200 | **18000** | 48600 | **68500** | - | - |

Cells in bold indicate homologous responses to vaccine strains. Values for all three assays are antibody titers.
<sup>‡</sup> This virus contained an egg adaptation, D187V.
<sup>\*</sup> The similar virus A/Hong Kong/45/2019 was used in place of A/Minnesota/41/2019 for the FRNT assay
<sup>†</sup> The similar virus A/Cambodia/e0826360/2020 was used in place of A/Tasmania/503/2020 for the FRNT assay

The results in year 2 showed contrasting patterns, in that first-time vaccinees mounted slightly stronger responses to the prior-season vaccine strains than to the current-season vaccine strains. First-time vaccinees in year 2 had slightly higher day 30 GMTs to A(H1N1) A/Wisconsin/588/2019 (the vaccine strain) than did first-time vaccinees receiving A/Hawaii/70/2019 the previous year (60.6 vs. 40), but first-time vaccinees in year 2 had approximately threefold higher titers to the previous year’s strain, A/Hawaii/70/2019, than to the strain with which they were vaccinated (180.6 vs. 60.6, respectively) (Table 3). Similarly, first-time vaccinees in year 2 mounted approximately 1.5-fold higher titers to the previous year’s H3N2 strain than they did to the H3N2 strain in the vaccine (67.2 to A/Hong Kong/45/2019 and 45.6 to A/Cambodia/e0826360/2020). Similar to the patterns for H1N1, first-time vaccinees had slightly higher absolute post-vaccination titers to the vaccine strain (A/Cambodia/e0826360/2020) compared to individuals vaccinated with A/Hong Kong/45/2019 the previous year (titers of 45.6 vs. 34.7), showing the strain update still led to increased titers to the intended strain.

That first-time vaccinees in year 2 mounted higher titers to the prior year’s influenza A vaccine strains than to the current vaccine strains might reflect a strong influence of prior immunity, but it could also reflect differences in receptor avidity between the strains. Viruses that bind to cells with lower avidity are more easily neutralized by antibodies compared to viruses that bind with higher avidity, irrespective of antigenic differences [30]. To address this, we measured total HA-specific IgG antibody responses by ELISAs which are not affected by differences in viral receptor binding avidities, to measure antibody binding to the influenza A(H1N1) and A(H3N2) vaccine strains, with results shown in Figure 3 and Table 3. We found that the year 2, geometric mean day 30 antibody levels to the current vaccine strain were similar to or slightly higher than those to the previous vaccine strain in first-time vaccinees, suggesting no disproportionate boosting of prior immunity targeting past strains. That there were no statistically significant differences in the geometric mean post-vaccination antibody levels measured by ELISA between first-time vaccinees in years 1 and 2 suggests the previously reported low HAI and FRNT titers to A/Wisconsin/588/2019 and A/Cambodia/e0826360/2020 thus likely arise from differences in the receptor avidity. The antigenic distances implied by the ELISA data are also smaller and do not show the same asymmetry as before. For instance, the amount of antibody reactive to A/Wisconsin/588/2019 after vaccination with A/Hawaii/70/2019 (group 5, year 1, day 30) was similar to the amount of antibody reactive to A/Hawaii/70/2019 after vaccination with A/Wisconsin/588/2019 (group 4, year 2, day 30) (Table 3). Finally, consistent with the data from the HAI and FRNT assays, there were no statistically significant differences in absolute post-vaccination antibody levels measured by ELISA between first-time and repeat vaccinees in year 2.

**Figure 3.**
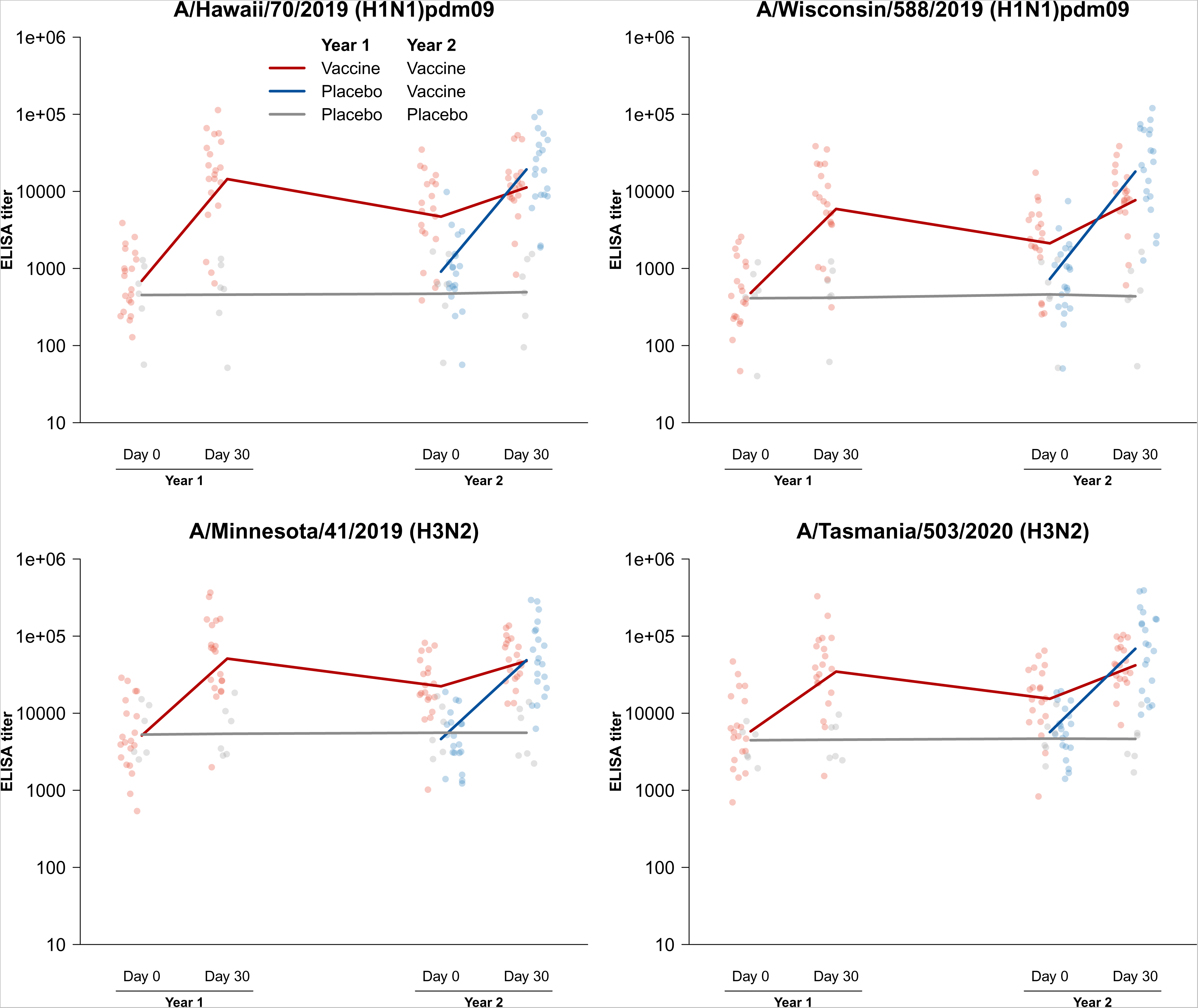
Antibody levels at various timepoints measured by enzyme-linked immunosorbent assay for influenza A(H1N1) and A(H3N2). Measured titers are plotted at 0 and 30 days post-vaccination in years 1 and 2, and lines represent the geometric mean antibody levels at each timepoint. Data from group 5, receiving vaccine in both years, are shown in red. Data from group 4, receiving placebo in year 1 and vaccine in year 2, are shown in blue. Data from the other groups, receiving placebo in both years, are shown in gray.

## DISCUSSION

In the second year of this five-year trial we identified reduced fold-rises in HAI titers after repeat vaccination for influenza A(H1N1), B/Victoria and B/Yamagata (Figure 2). However, post-vaccination GMTs were similar in repeat vaccinees and first-time vaccinees, indicating that these reduced responses likely would not hinder overall protection assuming that the post-vaccination HAI titer correlates with protection for Flublok [31]. During the study period, public health measures used to contain COVID-19 also prevented the community circulation of influenza [23], and our analysis of antibody titers is therefore unaffected by any potential differences in incidence of influenza virus infections in vaccine versus placebo recipients in the first year of the study that could have occurred if influenza had been circulating.

In year 2 of our study we found that repeat and first-time vaccinees also had similar post-vaccination GMTs to A(H3N2) measured by FRNT (Figure 2), indicative of similar levels of clinical protection. However, there was no substantial blunting of the fold rises to A(H3N2) in repeat vaccinees. Both groups started with low GMTs to A/Cambodia/e0826360/2020 (H3N2) in year 2 and increased those GMTs approximately 4-fold by 30 days post-vaccination (Figure 2, Appendix Table 4). Our ELISA analyses measuring direct antibody binding suggest that the observed differences in HAI and FRNT titers between groups could be due to differences in receptor binding avidities of the viral strains used in our studies.

Both the A(H1N1) and A(H3N2) vaccine strains were updated between years 1 and 2 of the study, raising opportunities to investigate the cross-reactivity of antibody titers induced by vaccination. The 2020/21 vaccine induced HAI titers to A(H1N1) and FRNT titers to A(H3N2) that were approximately 3-fold lower to the strains used in the 2021/22 vaccine. The suggested benefits of a vaccine update were evidenced by slightly higher titers to those strains among first-time vaccinees the following season compared to titers of vaccinees from the 2020/21 season. However, first-time vaccinees in the 2021/22 season did not have titers that were three-fold lower to strains from the 2020/21 season, as might be expected after vaccination in naïve animals or associated cartographic methods. Instead, individuals vaccinated in 2021/22 had approximately 3-fold and approximately 1.5-fold higher post-vaccination GMTs to prior season’s A(H1N1) and A(H3N2) vaccine strains. Measuring anti-HA antibody responses with ELISA implied even higher cross-reactivity and limited antigenic distances between viral strains used in successive years. These results suggest a need to interrogate antibody responses through multiple measures and to understand how each relates to protection from infection, disease and transmission.

Our study has a number of limitations. To preserve specimens for later longitudinal analyses, we have analyzed a subset of participants from the original study, and thus are limited in the effect sizes we can detect in the present analysis. Future analyses might yield different results. While the strains used in our serologic assays did not have 100% sequence identity with the vaccine strains, the minor genetic differences likely did not affect our major conclusions. Additionally, although the ELISA data suggest that antibody responses were comparable in magnitude between year 1 and 2 influenza A vaccine strains, at present we cannot conclude whether the contrasting HAI and FRNT results reflect strain-specific differences in receptor binding avidity, neutralization potential, and/or the relative immunogenicity of the stalk and other epitopes that are poorly measured by HAI. Understanding the roles of these factors is an important area for further work [32].

In conclusion, our randomized clinical trial of repeat influenza vaccination has found that repeat vaccination can be associated with reduced fold changes in antibody titers, but our preliminary results show no evidence of reduced post-vaccination mean titers, consistent with similar levels of protection after vaccination regardless of vaccination history. The apparently low immunogenicity of influenza A vaccine strains in 2021/22 relative to the prior year may reflect increases in receptor avidity of updated vaccine strains and deserves further study.

## Data Availability

All data produced in the present study are available upon reasonable request to the authors

## ACKNOWLEDGMENTS

The authors thank Suk Yee Chan, Tin Kin Chau, Trushar Jeevan, Pat Kaewpreedee, Ida Lam, Erica Lau, Kelly Lee, Maggie Lo, Charlie Man, Tiffany Ng, Yammy Ng, Yvonne Ng, Eunice Shiu, Lewis Siu, Tiffany Tavares, Ivan Tsai, Lilly Wang, Angel Wong, Irene Wong, Miyuki Wong, Phebe Yeung, and Zoe Xiao for technical support, and Julie Au and Ada Lee for administrative support.

