## Supplementary Materials for "Preliminary findings from the Dynamics of the Immune Responses to Repeat Influenza Vaccination Exposures (DRIVE I) Study: a Randomized Controlled Trial"


| | Page |
|---|---|
| 1. Summary of study design | 2 |
| 2. Vaccine strains used in the quadrivalent recombinant influenza vaccine | 5 |
| 3. Additional information on laboratory methods | 6 |
| 4. Reasons provided for participant withdrawal | 9 |
| 5. Selection of a subset of participants for serological analysis | 10 |
| 6. Comparison of geometric mean antibody titers | 11 |
| 7. Additional serologic testing comparing HAI titers against influenza A(H1N1) detected using egg-based or cell-based virus | 12 |
| 8. References | 13 |

### 1. Summary of study design

The study design of the DRIVE-I study is shown in Appendix Figure 1 below, with number of participants receiving the intervention in each group in years 1 and 2 shown in the figure.

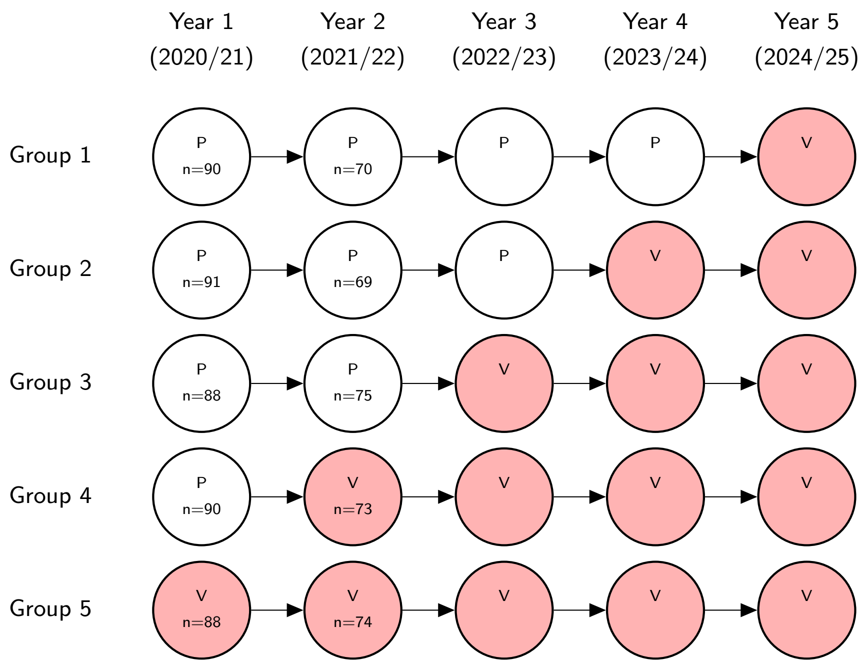


Appendix Figure 1. Randomization scheme of the DRIVE-I study. Participants were randomized to one of the five groups at the start of year 1. “P” indicates receipt of 0.5ml saline placebo, “V” indicates receipt of vaccination with FluBlok (Sanofi Pasteur). Actual sample sizes for each group in years 1 and 2 are shown inside the circles.

Participants were enrolled from the general community in Hong Kong, with study advertisements distributed via institutions (such as schools and universities), organizations (such as professional associations), and local community centers, and through mass promotion efforts including mass mailing to residential estates, advertisements in newspapers and public transport, social media platforms (such as Facebook), bulk emails, and invitation to and referrals from members of previous studies.

Potential participants were eligible for this study if they meet the following inclusion criteria and meet none of the exclusion criteria:

Inclusion criteria:

1. Aged 18-45 years at enrolment.

2. Capable of providing informed consent.
3. Resident in Hong Kong in the coming 2 years.

Exclusion criteria:

4. Vaccinated against influenza in the preceding 24 months.
5. Included in one of the priority groups to receive influenza vaccination in Hong Kong (priority groups include pregnant women, long-stay residents of institutions for persons with disability, persons with chronic medical problems (chronic cardiovascular, lung, metabolic or kidney diseases, obesity (body mass index 30 or above) and chronic neurological condition[1]), healthcare workers or persons working in poultry, pig farming or pig slaughtering industry).
6. With diagnosed medical conditions related to their immune system.
7. Currently taking medication for any condition that impairs immune system.
8. Individuals who report medical conditions not suitable to receive inactivated influenza vaccines, such as:
   - Severe allergic reaction (e.g., anaphylaxis) after previous dose of any influenza vaccine; or to a vaccine component;
   - Moderate or severe acute illness with or without fever after any previous influenza vaccination; or
   - A history of Guillain-Barré syndrome (GBS) within 6 weeks of previous influenza vaccination.
9. Individuals, who report medical conditions not suitable to receive intramuscular injection, such as
   - bleeding disorders
   - habitually taking anticoagulants (with the exception of antiplatelets such as aspirin).
10. Individuals who have any medical conditions not suitable to receive inactivated influenza vaccines as determined by a clinician.

[1] Information on the locally applied criteria for "high-risk conditions" can be found via https://www.chp.gov.hk/en/features/100764.html

We monitor pregnancy status in female participants throughout the study, and female participants will be unenrolled from the study if they are pregnant at a scheduled study vaccination and therefore encouraged to receive influenza vaccination as recommended by the Hong Kong government. We will similarly monitor job type and self-reported health conditions and immune conditions throughout the study and withdraw patients when they do not meet those inclusion criteria.

### 2. Vaccine strains used in the quadrivalent recombinant influenza vaccine

Appendix Table 1 below shows the strains used in the vaccine used in our study i.e. the northern hemisphere formulation of quadrivalent recombinant influenza vaccine (Flublok, Sanofi Pasteur) in 2020/21 and 2021/22.

Appendix Table 1. Influenza vaccine strains used in Flublok (Sanofi Pasteur) in the first two years of the DRIVE-1 study.

| | Year 1 (2020/21 Northern Hemisphere) | Year 2 (2021/22 Northern Hemisphere) |
|---|---|---|
| A(H1N1) | A/Hawaii/70/2019 | A/Wisconsin/588/2019 |
| A(H3N2) | A/Minnesota/41/2019 (an A/Hong Kong/45/2019-like virus) | A/Tasmania/503/2020 (an A/Cambodia/e0826360/2020-like virus) |
| B/Victoria | B/Washington/02/2019 | B/Washington/02/2019 |
| B/Yamagata | B/Phuket/3073/2013 | B/Phuket/3073/2013 |

### 3. Additional information on laboratory methods

Serum samples were treated with receptor destroying enzyme (RDE) and tested by HAI against the vaccine strains using a standard protocol [1]. For influenza A(H1N1), the vaccine strains were A/Hawaii/70/2019 in 2020/21 (GISAID Accession # EPI397028) and A/Wisconsin/588/2019 (GISAID Accession # EPI404460) in 2021/22. The stock for A/Hawaii/70/2019 were propagated in MDCK cells while A/Wisconsin/588/2019 were propagated in eggs once, due to the inability to obtain sufficient volume of cell-grown virus stock. A single mutation D204V was found in the egg-grown stock. A comparison of antigenicity using 22 serum samples from five participants showed that assay with the egg-grown stock had slightly higher sensitivity that yielded 2-fold higher titers in 50% of the samples but otherwise showed good correlation with cell-grown stock (Appendix Figure 2). For influenza B, ether-treated egg-grown antigens were used, and the vaccine strains were B/Washington/02/2019 (GISAID Accession # EPI347829) and B/Phuket/3073/2014 (GISAID Accession # EPI168822) in both years. Antibody titers were recorded as the reciprocal serum dilution in which the viral hemagglutination of 0.5% turkey red blood cells was inhibited. The original virus strains were received from the laboratory of Dr. Richard Webby at St. Jude Children's Research Hospital (Memphis, TN).

We used a focus reduction neutralization test (FRNT) for the two influenza A(H3N2) vaccine strains A/Hong Kong/45/2019 in 2020/21 and A/Cambodia/e0826360/2020 in 2021/22. A/Hong Kong/45/2019 HA (GISAID Accession # EPI1397376) is identical at the amino acid level to A/Minnesota/41/2019 HA (GISAID Accession # EPI1487157) included in the Flublok for 2020/21 influenza season. However, A/Cambodia/e0826360/2020 HA (GISAID Accession # EPI1837753) differs by a single amino acid substitution (N171K at antigenic site D) from A/Tasmania/503/2020 HA (GISAID Accession # EPI1759269) included in the

Flublok for 2021/22 influenza season. Both influenza A(H3N2) virus strains were generated by reverse genetics using A/Puerto Rico/8/1934 internal genes [2]. FRNT assays were completed as previously described [3]. Briefly, serum samples were treated with receptor destroying enzyme (RDE) (Denka-Seiken) for two hours at 37°C and then inactivated for 30 min at 56°C. Then, RDE-treated serum samples were 2-fold serially diluted in a 96-well plate using serum-free MEM and mixed with ~300 FFUs of virus per well. After 1 h incubation at room temperature, virus–serum mixtures were transferred to confluent monolayers of MDCK-SIAT1 cells. After 1 h incubation at 37 °C in 5% $CO_2$, cells were washed with serum-free MEM and overlaid with MEM supplemented with 5 mM HEPES buffer, 50 μg/mL gentamycin-sulfate, and 1.25% Avicel. Plates were incubated for 18 h at 37°C in 5% $CO_2$ and then fixed with 4% paraformaldehyde for 45 min at 4°C. Fixed cells were permeabilized with 0.5% Triton X-100 for 7 min, and then blocked for 45 min with 5% milk in PBS. Foci were stained for 1 h with a mouse anti-influenza A nucleoprotein antibody (clone IC5-1B7) followed by 1 h incubation with a peroxidase-conjugated rat anti-mouse kappa antibody diluted in blocking buffer. Plates were washed 5 times with distilled water after blocking, primary, and secondary steps. TrueBlue TMB substrate (SeraCare) was added to plates and incubated in the dark for 30 min for foci development. Substrate was flicked out of wells, and plates were thoroughly dried before foci visualization and quantification on an ELISpot reader. Serum antibody titers were recorded as the greatest dilution that neutralized ≥90% of the virus, also referred to as $FRNT_{90}$ titers.

To estimate total binding antibody of specific IgG responses, Enzyme Linked Immunosorbent Assays (ELISAs) were performed with recombinant HA (rHA) of the A(H1N1) vaccine strains A/Hawaii/70/2019 in 2020/21 and A/Wisconsin/588/2019, and the A(H3N2) vaccine strains A/Minnesota/41/2019 and A/Tasmania/503/2020 (Appendix Table 1). The HA stalk monoclonal antibody CR9114 was used as an internal control on each plate. Expression and purification of rHAs and CR9114 was performed as previously described [4]. To perform the assay, Immunlon 4HBX 96-well plates were coated with 1 ug/ml rHA overnight at 4°C. Plates were washed with PBS + 0.1% Tween-20, then blocked with PBS + 0.1% Tween-20, 0.5% milk, and 3% goat serum. Serum samples were serially diluted 4-fold starting at 1:100 and CR9114 was serially diluted 2-fold starting at 1 ug/ml, both in blocking buffer. After washing plates, serum and antibody dilutions were added and incubated for 2 hr. Plates were washed and horseradish peroxidase-conjugated anti-human IgG secondary antibody (Jackson, 109-036-098; diluted 1:5000) was added and incubated for 1 hr. After washing, plates were developed with SureBlue TMB peroxidase substrate (SeraCare) for 5 min before quenching with 250 mM hydrochloric acid. Plates were read at 450 nm using a SpectraMax 190 microplate reader (Molecular Devices, San Jose, CA). Antibody titers were recorded as reciprocal endpoint dilutions normalized relative to the CR9114 control for each plate.

### 4. Withdrawal from study

Some participants withdrew from the trial during the first year of follow-up and did not receive vaccination in the second year. The reasons for withdrawal provided by participants are shown in Appendix Table 2.

Appendix Table 2. Reasons for withdrawal during the first year of follow-up among participants in DRIVE-1.

| Reason for withdrawal | Group 1 | Group 2 | Group 3 | Group 4 | Group 5 |
|---|---|---|---|---|---|
| Lost contact e.g. changed mobile telephone number without informing us | 12 | 6 | 5 | 5 | 7 |
| Unwilling to continue in study | 5 | 10 | 4 | 10 | 5 |
| Moved away from Hong Kong | 1 | 3 | 3 | 1 | 1 |
| Actual/planned pregnancy | 1 | 0 | 1 | 0 | 1 |
| Have received the influenza vaccine outside of the study | 1 | 2 | 0 | 0 | 0 |
| Worsening health | 0 | 1 | 0 | 1 | 0 |
| Adverse effects | 0 | 0 | 0 | 0 | 0 |
| Other reasons/Unknown | 0 | 0 | 0 | 0 | 0 |
| **Total** | **20** | **22** | **13** | **17** | **14** |

### 5. Selection of a subset of participants for serological analysis

We selected a subset of participants with complete sera from five timepoints, namely year 1 days 0, 30 and 182, and year 2 days 0 and 30. We selected 5 participants at random from each of groups 1-3 (receiving placebo in both years), and 40 participants at random from each of groups 4 and 5. The characteristics of these 95 participants are compared between groups in Appendix Table 3.

Appendix Table 3. Characteristics of the subset of 95 participants selected for serological analysis

| **Characteristic** | **Randomized allocation to receipt of placebo (P) or influenza vaccination (V) in the first two years** | | | **p-value*** |
|---|---|---|---|---|
| | **Groups 1-3 P-P (n=15)** | **Group 4 P-V (n=40)** | **Group 5 V-V (n=40)** | |
| Age at randomization | | | | |
| 18-25 years | 3 (20%) | 10 (25%) | 8 (20%) | |
| 26-35 years | 5 (33%) | 15 (38%) | 14 (35%) | |
| 36-45 years | 7 (47%) | 15 (38%) | 18 (45%) | 0.96 |
| Male sex | 8 (53%) | 23 (58%) | 17 (43%) | 0.44 |
| Ever received influenza vaccination | 3 (20%) | 4 (10%) | 4 (10%) | 0.52 |

*p-value using Fisher's exact test

### 6. Comparison of geometric mean antibody titers

Appendix Table 4. Geometric mean antibody titers against influenza A(H1N1), A(H3N2) and B in each group at various timepoints. Antibody responses were measured by hemagglutination inhibition assay for influenza A(H1N1) and B, and by focus reduction neutralization test for influenza A(H3N2).

| Group | Strain | Year 1 | | | Year 2 | |
|---|---|---|---|---|---|---|
| | | Day 0 | Day 30 | Day 182 | Day 0 | Day 30 |
| 1, 2 and 3 combined (placebo-placebo) (n=15) | A/Hawaii/70/2019 (H1N1) | 7.6 | 7.6 | 7.6 | 7.6 | 7.6 |
| | A/Wisconsin/588/2019 (H1N1) ‡ | 7.9 | 11 | 7.6 | 6.6 | 7.9 |
| | A/Minnesota/41/2019 (H3N2)* | 14 | 14 | 15 | 14 | 15 |
| | A/Tasmania/503/2020 (H3N2)† | 12 | 11 | 12 | 11 | 11 |
| | B/Washington/02/2019 (B/Victoria) | 17 | 17 | 18 | 16 | 17 |
| | B/Phuket/3073/2013 (B/Yamagata) | 35 | 40 | 46 | 53 | 50 |
| 4 (placebo-vaccine) (n=40) | A/Hawaii/70/2019 (H1N1) | 8.1 | 8.3 | 8.3 | 8.7 | 180 |
| | A/Wisconsin/588/2019 (H1N1) ‡ | 7.4 | 6.4 | 6.4 | 6.7 | 61 |
| | A/Minnesota/41/2019 (H3N2)* | 11 | 11 | 11 | 11 | 67 |
| | A/Tasmania/503/2020 (H3N2)† | 10 | 10 | 10 | 10 | 46 |
| | B/Washington/02/2019 (B/Victoria) | 13 | 13 | 12 | 11 | 72 |
| | B/Phuket/3073/2013 (B/Yamagata) | 39 | 36 | 39 | 50 | 230 |
| 5 (vaccine-vaccine) (n=40) | A/Hawaii/70/2019 (H1N1) | 6.3 | 130 | 56 | 46 | 84 |
| | A/Wisconsin/588/2019 (H1N1) ‡ | 6.7 | 38 | 21 | 17 | 44 |
| | A/Minnesota/41/2019 (H3N2)* | 12 | 107 | 57 | 49 | 130 |
| | A/Tasmania/503/2020 (H3N2)† | 11 | 35 | 20 | 18 | 65 |
| | B/Washington/02/2019 (B/Victoria) | 9.3 | 62 | 37 | 29 | 49 |
| | B/Phuket/3073/2013 (B/Yamagata) | 60 | 270 | 170 | 150 | 200 |

* The similar virus A/Hong Kong/45/2019 was used in place of A/Minnesota/41/2019 for the FRNT assay

† The similar virus A/Cambodia/e0826360/2020 was used in place of A/Tasmania/503/2020 for the FRNT assay

‡ This strain differs from the original by the D204V mutation.

7. **<u>Additional serologic testing comparing HAI titers against influenza A(H1N1) detected using egg-based or cell-based virus</u>**

We conducted additional testing to determine whether the A/Wisconsin/588/2019 strain with the egg-adapted mutation significantly differs from that of the cell-based virus in terms of HAI titer. We selected 22 samples for testing in parallel against egg-based (D204V) and cell-based viruses. The samples included five time points from four individuals i.e. year 1 days 0, 30 and 182 and year 2 days 0 and 30, plus an additional two samples from one individual at year 1 days 0 and 30. Appendix Figure 2 below shows the correlation of titers in the 22 samples.

- **Cell-based virus** - grown in humanized MDCK cell line with PR8 backbone; A/Wisconsin/588/2019 + PR8 (P+2) C2S2C1/C1 with an HA titer of 16.
- **Egg-based virus** – A/Wisconsin/588/2019 C2S2C1/E1 HA 256

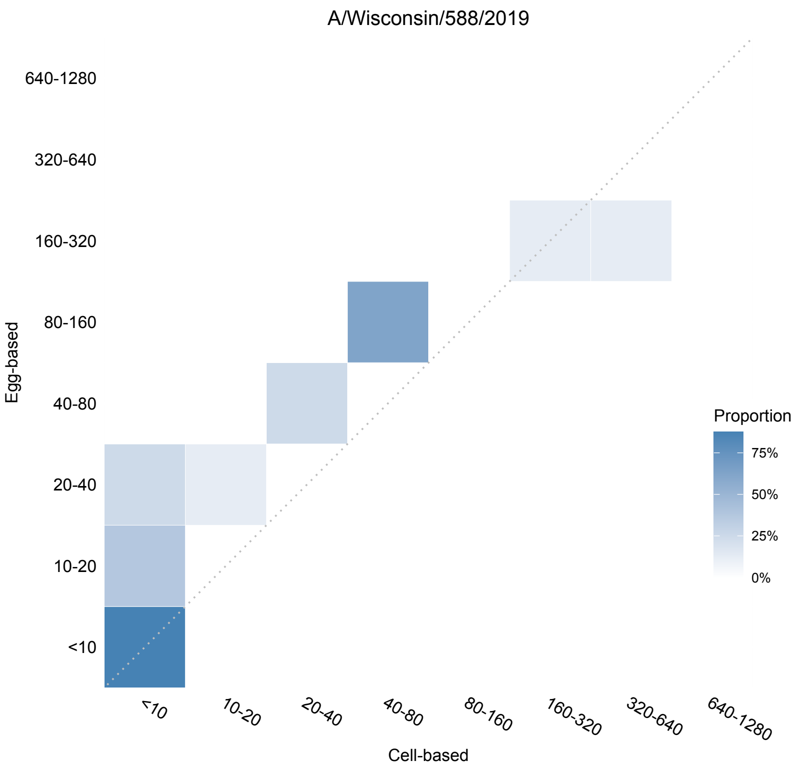


Appendix Figure 2: Antibody titers measured by the hemagglutination inhibition assay against cell-propagated A/Wisconsin/588/2019 and egg-propagated A/Wisconsin/588/2019. Egg-grown stock possess a D204V mutation. The dotted line is the diagonal i.e. identical titers against both strains.
